# Impact of COronaVirus Disease-2019 (COVID-19) pandemic on Haemodialysis care delivery pattern in Karnataka, India - a cross-sectional, questionnaire based survey

**DOI:** 10.1101/2020.07.25.20151225

**Authors:** Y J Anupama, Arvind Conjeevaram, A Ravindra Prabhu, Manjunath Doshetty, Sanjay Srinivasa, Venkatesh Moger

## Abstract

The COVID-19 pandemic has disrupted health care delivery globally. Patients on in-centre haemodialysis(HD) are particularly affected due to their multiple hospital visits and the need for uninterrupted care for their well-being and survival. We studied the impact of the pandemic and the national policy for pandemic control on the HD care delivery in Karnataka state in India in April 2020, when the first and second national lockdown were in place. An online, questionnaire based survey of dialysis facilities was conducted and the responses analysed. The questions were pertaining to the key areas such as changes in number of dialysis treatments, frequency, duration, expenses, transportation to and from dialysis units, impact on availability of consumables, effect on dialysis personnel and on machine maintenance. 62 centres participated. Median of dialysis treatments for the months of March and April 2020 were 695.5 and 650 respectively. Reduction in dialysis treatments was noted in 29(46.8%) facilities, decreased frequency reported by 60 centres. In at least 35(56.5%) centres, dialysis patients had to bear increased expenses. Cost and availability of dialysis consumables were affected in 40(64.5%) and 55(88.7%) centres respectively. Problems with transportation and movement restriction were the two key factors affecting both patients and dialysis facilities.This survey documents the collateral impact of COVID-19 on the vulnerable group of patients on HD, even when not affected by COVID. It identifies the key areas of challenges faced by the patients and the facilities and implores the care-providers for finding newer avenues for mitigation of the problems.

---

The coronavirus disease 2019 (COVID-19) pandemic has affected millions of people globally. While the direct effects of the virus are evident in the surging numbers of patients, it has also disrupted healthcare delivery in almost all countries. The surge in COVID 19 cases with resultant allocation of beds to such cases has often forced a decrease in admissions to non-COVID cases.^1^ Delivery of healthcare in diverse areas such as maternal and child health, tuberculosis, cancer treatment and dialysis have been affected in many countries.^2^ The patients on chronic haemodialysis form a unique subset of patients particularly affected by these changes in healthcare delivery.^3^ They have a compelling reason to come to the healthcare facility frequently, at least twice or thrice a week, which now became difficult due to many reasons.

The Government of India, imposed its first national lockdown on March 24th 2020 for a period of 21 days, with subsequent extensions through April and May.^4^ The resultant movement restrictions as well as lack of public transport has impacted patients on dialysis. In addition, several hospitals were designated COVID hospitals and the patients undergoing dialysis in those centres were reallocated to different hospitals. This has brought about numerous challenges both to patients on dialysis and dialysis facilities alike.

We studied the impact of these changes in the treatment of patients undergoing dialysis in Karnataka, a state in South India. The state has a population of 64 million and has 30 districts.^5^ The capital city is Bengaluru which is located in south Karnataka. There are over 200 dialysis facilities all over Karnataka, with nearly 50% located in Bengaluru. The remaining facilities are distributed in the rest of the state, in smaller cities and towns. Most of the centres outside Bengaluru depend on supply of dialysis consumables from Bengaluru. The facilities outside Bengaluru generally have more patients from rural areas compared to centres in Bengaluru, which is a metropolitan city. We also examined whether there were significant differences between the centres in and outside Bengaluru (hereafter designated as Metro and Non-Metro centres).

## Materials and methods

We invited nephrologists from all over Karnataka to participate in a questionnaire based survey of changes in dialysis practice pattern during the month of April 2020 when the national lockdown was in force. The questionnaire was sent out to all (nearly 200) as a Google Form and replies collected. The questions were pertaining to eight key areas that could affect dialysis delivery. Dialysis numbers, frequency, duration, expenses, transportation to and from dialysis units, impact on availability of consumables, effect on dialysis personnel and on machine maintenance. Results were analysed using SPSS version 16(SPSS Inc., Chicago IL). Descriptive data was analysed as percentages to measure proportions and medians for continuous variables. Categorical variables were compared among Metro and Non-metro centres using chi square test. P< 0.05 was taken as significant.

## Results

In all, 62 centres participated, of whom 31(50%) were from Bengaluru (Metro centres) and another 50% from outside Bengaluru (Non –metro centres) (Figure 1).There was representation from nearly 15 districts. The key features of the centres are represented in Table 1. One response was collected from each centre, irrespective of the number of nephrologists in those centres. Nephrologists providing dialysis care in more than one facility had to provide one form per facility.

**Table 1:** Overview of participating centres.

| **Table 1: Overview of participating centres** |  |  |
| --- | --- | --- |
| **Centre Characteristics** | **Number** | **Percentage** |
| No of centres | 62 | 100 |
| Centres in Bengaluru | 31 | 50 |
| Centres outside Bengaluru | 31 | 50 |
| Type of Hospital |  |  |
| Government | 4 | 6.5 |
| Teaching Hospital | 13 | 21 |
| Private Hospital | 43 | 69.4 |
| Trust- based | 2 | 3.2 |
| Number of Nephrologists |  |  |
| 1 | 29 | 46.8 |
| 2 | 20 | 32.3 |
| 3 or more | 13 | 20.9 |
| Number of machines |  |  |
| <5 | 9 | 14.5 |
| 6-10 | 18 | 29 |
| 11-15 | 18 | 29 |
| 16-20 | 3 | 4.8 |
| 21-25 | 5 | 8.1 |
| >25 | 9 | 14.5 |

**Figure 1:**
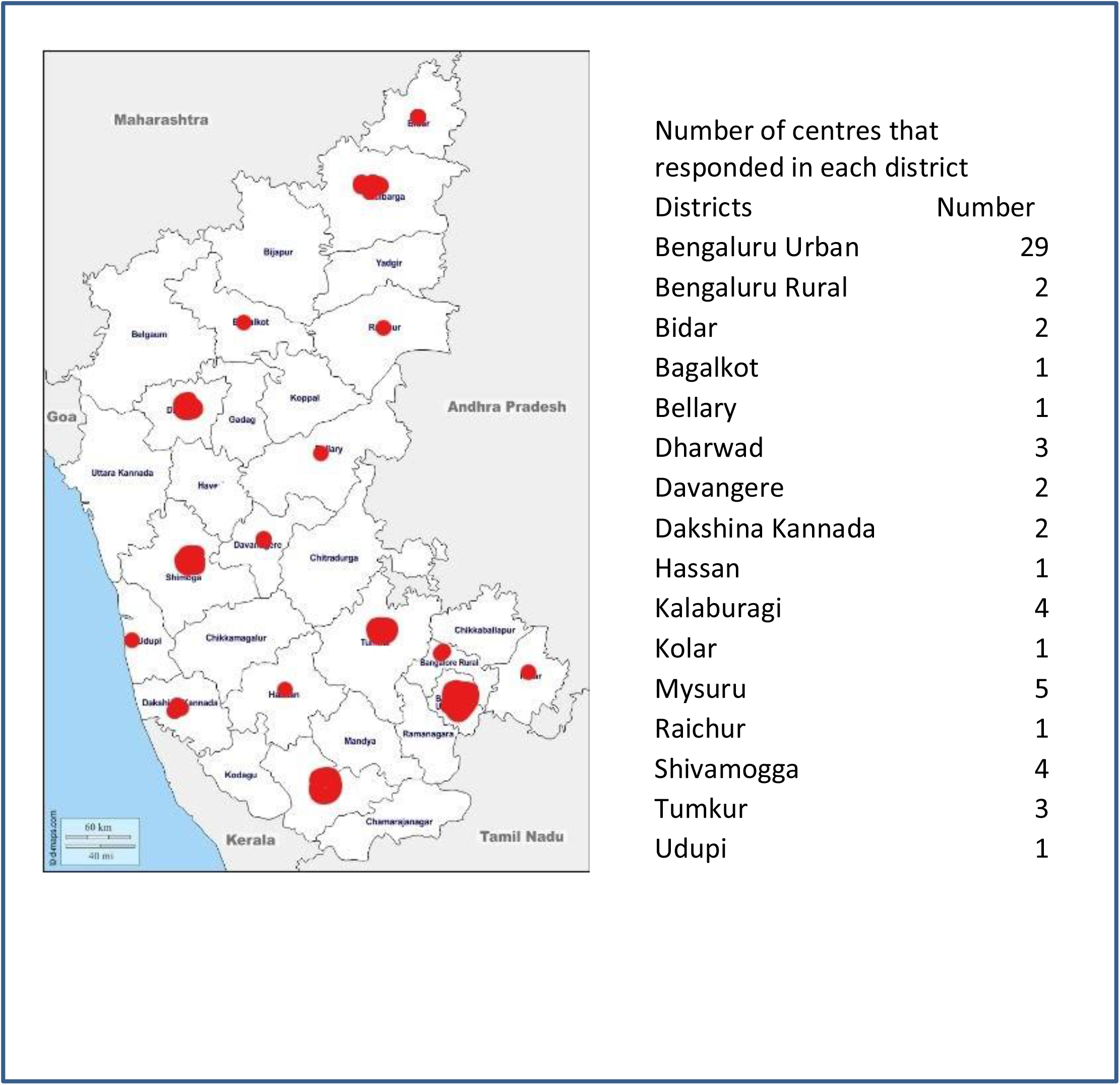
Depiction of centres contributing data and their location.

The median(Inter Quartile Range) of the dialysis treatments for the months of March 2020 and April 2020 were 695.5(353,999) and 650(317, 982) respectively. Corresponding median(IQR) for dialysis treatments for the month of April 2019 was 700(362,1027). There were changes in dialysis care delivery perceived both due to patient–related factors and logistic related issues (Table 2).

**Table 2:** Overview of factors affected at patient-level and at dialysis centres

|  |  |  |  |  |
| --- | --- | --- | --- | --- |
| Variable | All centres(n=62),<br>Number(%) | Metro<br>Centres<br>(n=31),<br>Number (%) | Non- metro<br>Centres( n=31),<br>Number(%) | p |
| **Patient-related challenges** |  |  |  |  |
| Dialysis Numbers |  |  |  |  |
| Decreased | 29(46.8) | 16(51.6) | 13(41.9) | **0.021** |
| Increased | 15(24.2) | 3(9.7) | 9(38.7) |  |
| No change | 18(29) | 12(38.7) | 6(19.4) |  |
| Decreased Dialysis duration | 4(6.5) | 1(3.2) | 3(9.7) | 0.301 |
| Percentage affected by reduced dialysis frequency |  |  |  |  |
| <10% | 43(69.4) | 23(74.2) | 20(64.5) | 0.367 |
| 11-20% | 8(12.9) | 3(9.6) | 5(16.1) |  |
| >20% | 9(14.5) | 3(9.6) | 6(19.3) |  |
| *Reasons for reduced frequency |  |  |  |  |
| Increased cost | 5(8.1) | 5(16.1) | 0(0.0) | 0.173 |
| Problems with transportation | 49(79.1) | 22(70.9) | 27(87.1) |  |
| Restriction of movement | 11(17.8) | 5(16.1) | 6(19.3) |  |
| Limited dialysis manpower | 7(11.2) | 5(16.1) | 2(6.4) |  |
| To conserve consumables | 2(3.2) | 1(3.2) | 1(3.2) |  |
| Increased dialysis expenses | 35(56.5) | 15(48.4) | 20(64.5) | 0.2 |
| Percentage affected by increased cost of transportation |  |  |  |  |
| <20% | 30(48.4) | 17(54.8) | 13(41.9) | **0.047** |
| 21-40% | 16(25.8) | 5(16.1) | 11(35.5) |  |
| >40% | 14(22.6) | 7(22.6) | 7(22.6) |  |
| **Logistic-related challenges** |  |  |  |  |
| *Non-availability of consumables(overall) | 55(88.7) | 27(87.1) | 28(90.3) | 0.79 |
| Dialyzers | 33(53.2) | 16(51.6) | 17(54.8) |  |
| Heparin | 33(53.2) | 14(45.2) | 21(67.7) |  |
| Dialysis concentrates | 23(37.1) | 11(35.5) | 12(38.7) |  |
| *Increased cost of consumables (overall) | 40(64.5) | 15(48.4) | 25(80.6) | **0.009** |
| Dialyzers | 24(38.7) | 12(38.7) | 12(38.7) |  |
| Heparin | 35(56.4) | 11(35.5) | 24(77.4) |  |
| *Dialysis personnel related issues | 45(72.5) | 23(74.2) | 22(71) | 0.708 |
| Absenteeism | 22(35.5) | 10(32.3) | 12(38.7) |  |
| Psychological issues | 27(43.5) | 13(41.9) | 14(45.2) |  |
| Difficulties with transportation | 28(45.2) | 16(51.6) | 12(38.7) |  |
| Number of centres increasing cost per sitting | 14(22.5) | 10(32.3) | 4(12.9) | 0.068 |
| Problems with maintenance of machines | 32(51.6) | 8(33.3) | 24(66.6) | **<0.001** |

\* More than one factor may be responsible (hence total exceeds 100% in places)

Patient-related domains: Overall 29(46.8%) centres reported a decrease in the number of dialysis treatments, while 18(29%)had no change and 15(24%) reported an increase. The decrease was mostly due to movement restriction due to lockdown(82.8%) and decrease in frequency of dialysis due to difficulties in transportation(75.9%). In 13.8% cases, the dialysis centre was not functioning to full capacity as the hospital had been converted to a COVID hospital. Conversely, 15 centres recorded an increase in dialysis treatments, mostly due to transfer of patients from government hospitals which had been converted to COVID. Of the 62 centres, only 4(6.5%) reported decrease in dialysis duration. There was a reported decrease in frequency of dialysis in 60 facilities. In 43(69%) centres, this was affecting less than 10% patients while in 9 (14.5%) centres, more than 20% patients were affected by the reduction in frequency. In a resource-constrained setting, with the national lockdown there arose difficulties in transportation. Most patients, who were dependent on public transport to travel to and from the centre, now had to hire private transport and this added to the transportation expenses borne by the individual patient. In all, 35(56.5%) centres reported that the patients had to face the burden of increased expenses and at least in 14(40%) of these, more than 40% patients were affected by this. This was found to be more profound in facilities outside Bengaluru. The out-of–pocket expenses that were borne by patients went up by 50-200% in most instances.

Logistic related challenges: Dialysis facilities too had to face many challenges. The dialysis consumables were in short supply. The sudden cessation of transport impacted the supplies as well and as many as 55(88.7%)centres, especially outside Bengaluru, faced shortages. Most affected were supplies of heparin and dialyzers with shortages affecting more than 30% centres. The supply of dialysis concentrates was affected in 23(37.1%) centres. In all, 15(24%) centres faced shortages of all consumables. The cost of consumables too increased and this affected 40(64.5%) of the centres. This in turn, led some care providers (n=14, 22.6%) to increase the charges of dialysis sittings. The increase in charges ranges from 90-600₹ with median being 170 INR (170₹) per sitting. The difficulties were also felt in the area of maintenance of machines with 32(52%) centres affected due to inability of machine service personnel to attend to the peripherally located centres. This problem was experienced more by non-metro centres (p<0.05).

In addition, questions were also asked to know the impact of the COVID19 on the general dialysis practice of nephrologists. In all, 16% felt that the non-COVID infections have decreased in their units, possibly as a result of improved hand hygiene practices of dialysis staff. Only 16% felt that the death rates have gone up while 74% were not sure of any change in death rates. The precautions taken in dialysis units to prevent Coronavirus infection may translate into better dialysis care in the ensuing months and at least 40% felt it had a positive impact on their infection control practice, while 32% felt that it did not impact their care and another 27% were not sure of any change.

## Discussion

In this cross-sectional survey of dialysis facilities across the state of Karnataka, India, we have recorded a spectrum of effects on haemodialysis care delivery due to COVID-19 pandemic and the national response to control the spread of infection. We could demonstrate a wide range of problems, with the most important being related to transportation problems and movement restrictions, affecting the patients and dialysis supply chain alike.

This survey pertained to the impact on dialysis centres during April 2020, at which the country was going through the first and the second nation-wide lockdown. In India, the majority of dialysis units are located in private hospitals.^6^ In many states with a heavy burden of COVID-19, governments have taken over management of a number of private hospitals and ordered temporary closure of dialysis units. In some instances, with the detection of a SARS-CoV-2 positive patient on dialysis, and several centres had to be sanitized, staff quarantined and patients sent to different units leading to a lot of disruption in dialysis care delivery^7^ In all such instances, patients are asked to go to other dialysis facilities, and there are instances of those dialysis facilities not being able to accommodate such patients, putting the patients in great misery.^8^ In Karnataka, however, the situation(at the time of writing this article) is less grim in comparison to other states like Maharashtra and Gujarat.^9^ The COVID-19 case burden in the state at the time of this survey was mild to moderate with confirmed cases being 565 and deaths, being 23 (as of April 30, 2020) as against national numbers of 36607 and 1229 for confirmed cases and deaths respectively.^10^ Hence the challenges faced by patients on HD in Karnataka was less profound than in the other states as can be seen by the nearly unchanged dialysis numbers. However, in response to COVID-19 pandemic, there were some changes made in the state with many Government hospitals in the state being designated COVID hospitals and consequently the patients on HD from these hospitals were relocated to many private hospitals. This was one main reason for the increased workload in at least 24% dialysis facilities. This stretched the already limited workforce and the functioning of machines which forced some centres to decrease dialysis frequency as well as duration, thereby affecting the dialysis quality.

One major effect of COVID lockdown was the movement restriction which affected patient travel to hospitals as well as supply of dialysis consumables. The patients experienced great hardship as they had to depend on private transport and hence spend more for transport during this period. There were also administrative hassles with patients being stopped in various checkposts by the police and they had to obtain special passes for travel. The patients undergoing dialysis in non-metro centres often come from remote areas and travel nearly 100-150 km for each dialysis. The increase in weekly dialysis –related expenses, was felt more profoundly by these patients. With the easing of lockdown towards, mid-May, many of these problems are getting mitigated.

It is clear from this survey, that multiple domains of care were affected during the current COVID-19 pandemic impacting the patients and the dialysis facilities alike. There were widespread problems, not only in HD services, but also overall Nephrology services during this period. Kidney transplantation numbers and inpatient services were also seriously affected. In April 2020, there was a 95% reduction in live kidney transplants and there were no deceased donor transplants done during this period.^11^ Inpatient general nephrology burden showed a drastic reduction as can be inferred from the kidney biopsy numbers. Kidney biopsies reported during the month of April 2020 (Native 62, Allograft 11) in one of the largest nephropathology units in the state, showed a precipitous decline of 84% compared to April 2019 (Native 401, Allograft 64).^12^ This disruption in nephrology services assumes significance because it occurred at a time when the overall burden of COVID-19 was low in Karnataka and also in India. Though the dialysis facilities faced the present challenge well with most of them successfully accommodating all the patients and providing dialysis at no extra cost, this pandemic has alerted us to the future possibility of an unprecedented disruption of dialysis care delivery and exposed the vulnerability of our existing care model. The lessons for the dialysis facilities should be to develop contingency measures for future challenges. They should foster self-help groups in association with non-governmental organizations to mitigate the unforeseen financial hardships that may be faced by the patients in such times. India is dependent on import of many of the dialysis consumables, including heparin. In a global crisis, the supply gets affected as countries close borders and the government should plan for such challenges in the future with development of indigenous capacity for the manufacture of dialysis consumables. Contingency planning and developing peripheral centres for storage or manufacture of medical supplies is important.

## Conclusions

This survey serves to identify the collateral impact of COVID 19 on the dialysis care delivery on patients on hospital HD, even when unaffected by COVID-19 and documents the challenges faced by the patients and the facilities. The dialysis population is a vulnerable group and must be supported by the Government during these periods of calamities. It implores the care-providers for finding newer avenues for mitigation of the problems and points out deficiencies which require to be addressed during such unexpected periods of uncertainty such as future pandemics or calamities.

## Data Availability

Available on request

## Acknowledgements

- We gratefully acknowledge the Nephrologists of Karnataka who participated in the survey and shared their data:
- Anil Mudda, Mudda Dialysis Centre Bidar
- Anitha A, Apollo Hospital, Bengaluru
- Anupama Y J, Nanjappa Hospital, Shivamogga
- Anvar Mohammed, Vijayanagar Institute of Medical Sciences, Bellary
- Arun K G, Narayana Multi Speciality Hospital, Mysuru
- Arvind Conjeevaram, The Bangalore Hospital, Bengaluru, Meenakshi People Tree Hospital,
- Bengaluru, Sagar Hospitals, Jayanagar, Bengaluru
- B H Santhosh Pai, Yenepoya Medical College, Mangaluru
- Basantkumar H S, People Tree Hospital, Bengaluru
- Chakko Korula Jacob, Bangalore Baptist, Hospital
- Dayanand A S, MaAx superspeciality Hospital, Shivamogga
- G K Prakash, Manipal Hospital, Malleswaram, Bengaluru
- Girisha Namagondlu, KMF Ranka Hospital, Bengaluru
- Guni Mahapatra, Command Hospital Air Force, Bengaluru
- H Mahabaleshwar Mayya, Sushruta Multi-Specialty Hospital, Hubballi
- Harshakumar H N, Sparsh Superspeciality Hospital, Bengaluru
- Isthiaque Ahmed, Narayana Healthcity, Bengaluru
- Kiran K K, JSS Hospital, Mysuru
- Kiranchandra Patro,NU Hospital, Padmanbhanagar,Bengaluru
- Kishan Aral, Trust Well Hospitals, Bengaluru
- Madhusudan H C, Mangala Hospital, Hassan
- Mahesh E, Ramaiah Memorial Hospital, Bengaluru
- Manjunath Doshetty, Chirayu Hospital, Kalaburagi, BRS Dialysis Unit, GIMS Hospital, Kalaburagi, Aarogya Hospital, Bidar
- Manjunath Kulkarni, Father Muller Medical College, Mangaluru
- Manjunath R, SDM Medical College and Hospital, Dharwad
- Manjunath S, SSNMC Super Specialty Hospital, Bengaluru
- Prashanth C Dheerendra, Dharma Kidney Care And Research Private Limited, Bengaluru
- Praveen Malavade, NU Hospital, Shivamogga
- Rajeev Agarwal, Bapuji Hospital, Davanagere
- Rashmi Shrinivasa,R L JalappaHospital, Kolar
- Ravi K R, Sahyadri Narayana Multispeciality Hospital,Shivamogga
- Ravindra Prabhu A, Kasturba Hospital, Manipal
- Renuka Satish, St John,s Medical College Hospital, Bengaluru
- S Venkataraman Raju, Raju Institute Of Nephrology And Urology, Bengaluru
- Sandeep Huilgol, Dr Huilgol’s Kidney Care, Bagalkot
- Sanjay Srinivasa, Sapthagiri Institute of Medical Science, Bengaluru, Suguna Hospital, Bengaluru,CV Raman Nagar Government Hospital, Bengaluru,Shirdi Sai Hospital, Bengaluru
- Santosh Kumar S, Chinmaya Mission Hospital, Bengaluru
- Sathishkumar M M, Vikram Hospital Pvt Ltd, Bengaluru
- Suma Raju, Narayana Multispeciality Hospital, HSR Layout, Bengaluru
- Swarna Shashi Bhaskar, Regal Hospital, Thanisandra, Bengaluru
- Topoti Mukherjee,Manipal Hospital Whitefield, Bengaluru
- Uthappa Puchimada M, Columbia Asia Hospital, Mysuru
- Venkatesh Moger, Tatwadarsha Hospital, Hubballi
- Vidyashankar, Aster CMI Hospital, Bengaluru, Rajarajeshwari Medial College Hospital,
- Bengaluru, ExcelCare Hospital, Bengaluru
- Vinay Badri, Rajiv Gandhi Superspeciality Hospital, Raichur
- Vipin Kaverappa, Sigma Hospital, Mysuru
- Vishwanath Patil, SSIMS&RC, SS Hospital.Davanagere
- Vishwanath S, Manipal Hopsital, Old Airport Road, Bengaluru
- Vivek Patil, VaatsalyaLife Hospital, Kalaburagi, United Hospital, Kalaburagi
- Yeswanth Gangaiah, Siddaganga Hospital, Tumkur, CG Kidney Care, Tumkur, THS Hospital,Tumkur

